# Association between Social Media Exposure and Self-Medication Practices: Public Health Implications in Punjab, Pakistan

**DOI:** 10.64898/2026.11.22.26368645

**Authors:** Ayesha Ashraf, Faiza Khalid, Mian Salman, Muhammad Sharjeel, Muhammad Zohaib Qasim, Muhammad Abu Baker Saddique

**Affiliations:** School of Pharmacy, University of Management and Technology, Lahore, Pakistan

**Keywords:** self-medication, social media, digital health, medicine advertisements, Punjab, Pakistan

## Abstract

**Background:** Social media has become an important source of health information, including information about medicines. Exposure to medicine-related content may affect decisions to use medicines without professional consultation.

**Objective:** To describe self-medication practices and social-media exposure among adults in Punjab, Pakistan, and explore associations between self-medication frequency and demographic, social-media-use, and attitudinal factors. Methods: A cross-sectional analytical survey was conducted among adults aged 18–60 years in Punjab between March and May 2026. The final analytic sample comprised 280 participants. A bilingual (Urdu/English) questionnaire assessed demographics, social-media use and health-content exposure, self-medication in the preceding six months, medicine categories, and attitudes. The questionnaire was reviewed by two clinical experts and piloted among 30 participants. Descriptive analyses, chi-square tests, and multinomial logistic regression were reported using IBM SPSS Statistics; significance was set at α=.05.

**Results:** Self-medication frequency was reported as Rarely by 117 participants (41.8%), Frequently by 130 (46.4%), and More Often by 33 (11.8%). Fever medicines were reported by 179 participants (63.9%) and gastrointestinal medicines by 96 (34.3%); multiple-response medicine categories were not mutually exclusive. Instagram was the most frequently selected social-media platform. Advertisement recall differed across self-medication frequency groups (χ^2^(22)=35.28, p=.036). Belief that medicines shown in advertisements/posts were safe was associated with self-medication frequency (χ^2^(4)=14.26, p=.007), while perceived risk was not (χ^2^(4)=1.587, p=.811). The multinomial model’s overall test was not statistically significant (χ^2^(40)=54.07, p=.068; Nagelkerke R^2^=.205). Likelihood-ratio tests identified associations for daily social-media-use duration (χ^2^(2)=13.085, p=.001), age group (χ^2^(6)=14.930, p=.021), and perceived advertisement safety (χ^2^(4)=10.899, p=.028).

**Conclusion:** Self-medication was commonly reported in this sample. Some social-media-use and advertising-attitude variables were associated with self-medication frequency, but the overall regression model was not statistically significant and the cross-sectional design cannot establish causality. Findings should be interpreted cautiously given the sample composition and reported data limitations.

## 1. Introduction

Self-medication is a public-health concern because inappropriate medicine selection, dosing, interactions, masking of symptoms, delayed care, and antimicrobial resistance may result. Social-media platforms have become widely used channels for health information and medicine-related promotional content, but the accuracy and reliability of such content can vary.

Pakistan’s growing digital environment makes it important to understand whether social-media exposure and attitudes toward online medicine promotion are associated with self-medication. Previous work in Pakistan has examined self-medication patterns and medicine use, while the relationship between social-media exposure, advertisement perceptions, and self-medication among adults in Punjab remains an area for further study.

This study aimed to describe self-medication practices and social-media exposure among adults in Punjab, Pakistan, and explore associations between self-medication frequency and demographic, social-media-use, and attitudinal factors.

## 2. Methods

### Study design and setting

A cross-sectional analytical survey was conducted among adults residing in Punjab, Pakistan. Data were collected from March to May 2026.

### Participants and eligibility

Eligible participants were aged 18–60 years, resided in Punjab, had smartphone and internet access, and consented to participate. The final analytic sample was 280. The thesis reports exclusion of incomplete or duplicate responses, individuals outside the age range, and individuals who did not consent.

### Questionnaire

A bilingual (Urdu/English), self-administered questionnaire covered socio-demographic characteristics; social-media use and health-related content exposure; self-medication during the preceding six months; medicine categories and information sources; and attitudes toward medicines promoted online. The questionnaire was reviewed by two clinical experts and piloted among 30 participants. The Urdu version was translated and back-translated.

### Sampling and data collection

The thesis describes recruitment through online channels and community-based approaches, including pharmacies and neighbourhood settings. The questionnaire was distributed through Google Forms and also administered through questionnaire-based interviews. No monetary incentives were offered.

### Variables

The outcome was self-reported self-medication frequency, categorised as Rarely, Frequently, or More Often. Exposures and covariates included daily social-media use, age group, gender, education, occupation, and attitudes toward medicine advertisements and online health information.

### Ethical considerations

The thesis acknowledges approval by the Research Ethics and Support Committee (RESC), University of Management and Technology, Lahore, Pakistan, with reference number RE-42-2026. It reports informed consent from participants. The exact approval document and reference number should be checked before submission.

### Statistical analysis

Descriptive frequencies and percentages were calculated. Multiple-response analyses described medicine categories, platforms, and advertisement recall. Associations were examined using chi-square tests and Fisher’s exact test where appropriate. Multinomial logistic regression (NOMREG) was reported, with “More Often” as the reference outcome category. Statistical significance was set at α=.05. Reported values should be checked against the original statistical output before submission.

## 3. Results

### Participant characteristics

The sample included 280 participants. Most were aged 18–25 years (206; 73.6%); 138 (49.3%) were male and 142 (50.7%) female. University-level education was reported by 199 (71.1%). Students/dependants accounted for 158 (56.4%) participants.

### Social-media use

Daily use was 4–6 hours for 102 participants (36.4%), 2–3 hours for 95 (33.9%), over 6 hours for 53 (18.9%), and under 1 hour for 26 (9.3%). A total of 139 participants reported following people who share medical journeys. Instagram was the most frequently selected platform (189 responses), followed by YouTube (92), Facebook (72), Google (57), and television advertisements (18).

### Self-medication and medicines

Self-medication frequency was Rarely 117 (41.8%), Frequently 130 (46.4%), and More Often 33 (11.8%). Fever medicines were reported by 179 participants (63.9%), gastrointestinal medicines by 96 (34.3%), beauty/cosmetic products by 57 (20.4%), energy boosters/supplements by 48 (17.1%), skin-related medicines by 45 (16.1%), antibiotics by 34 (12.1%), joint-pain medicines by 31 (11.1%), mood/anxiety/depression medicines by 21 (7.5%), sleep medicines by 15 (5.4%), PCOS medicines by 9 (3.2%), heart medicines by 9 (3.2%), and sexual-problem medicines by 5 (1.8%). Multiple-response categories were not mutually exclusive.

### Advertisement recall and attitudes

Hair-problem advertisements were most frequently recalled (182 responses), followed by weight-loss advertisements. Advertisement recall differed across self-medication frequency groups (χ^2^(22)=35.28, p=.036); the thesis notes small expected cell counts and advises caution. Belief that medicines shown in advertisements/posts were safe was associated with self-medication frequency (χ^2^(4)=14.26, p=.007; Fisher’s exact p=.006). Perceived risks of medication use from social media were not associated (χ^2^(4)=1.587, p=.811).

### Multinomial regression

The overall model test was χ^2^(40)=54.07, p=.068, with Nagelkerke R^2^=.205. Likelihood-ratio tests indicated associations for daily social-media-use duration (χ^2^(2)=13.085, p=.001), age group (χ^2^(6)=14.930, p=.021), and perceived advertisement safety (χ^2^(4)=10.899, p=.028). For the Rarely versus More Often comparison, daily social-media use had B=−0.940, p=.005, OR=0.391 (95% CI 0.202–0.756). The reported age comparison (18–25 versus 45– 60) had B=4.901, p=.008, OR=134.42 (95% CI 3.54–5109.55), with a very wide confidence interval.

**Table 1.** Participant characteristics (N=280)

| Characteristic | Category | n | % |
|---|---|---|---|
| Age | 18–25 | 206 | 73.6 |
|  | 25–35 | 45 | 16.1 |
|  | 35–45 | 22 | 7.9 |
|  | 45–60 | 7 | 2.5 |
| Gender | Male | 138 | 49.3 |
|  | Female | 142 | 50.7 |
| Education | School | 19 | 6.8 |
|  | College | 62 | 22.1 |
|  | University and above | 199 | 71.1 |
| Occupation | Private sector | 47 | 16.8 |
|  | Self-employed | 43 | 15.4 |
|  | Unemployed | 22 | 7.9 |
|  | Student/dependent | 158 | 56.4 |
|  | Government employee | 10 | 3.6 |

**Table 2.**
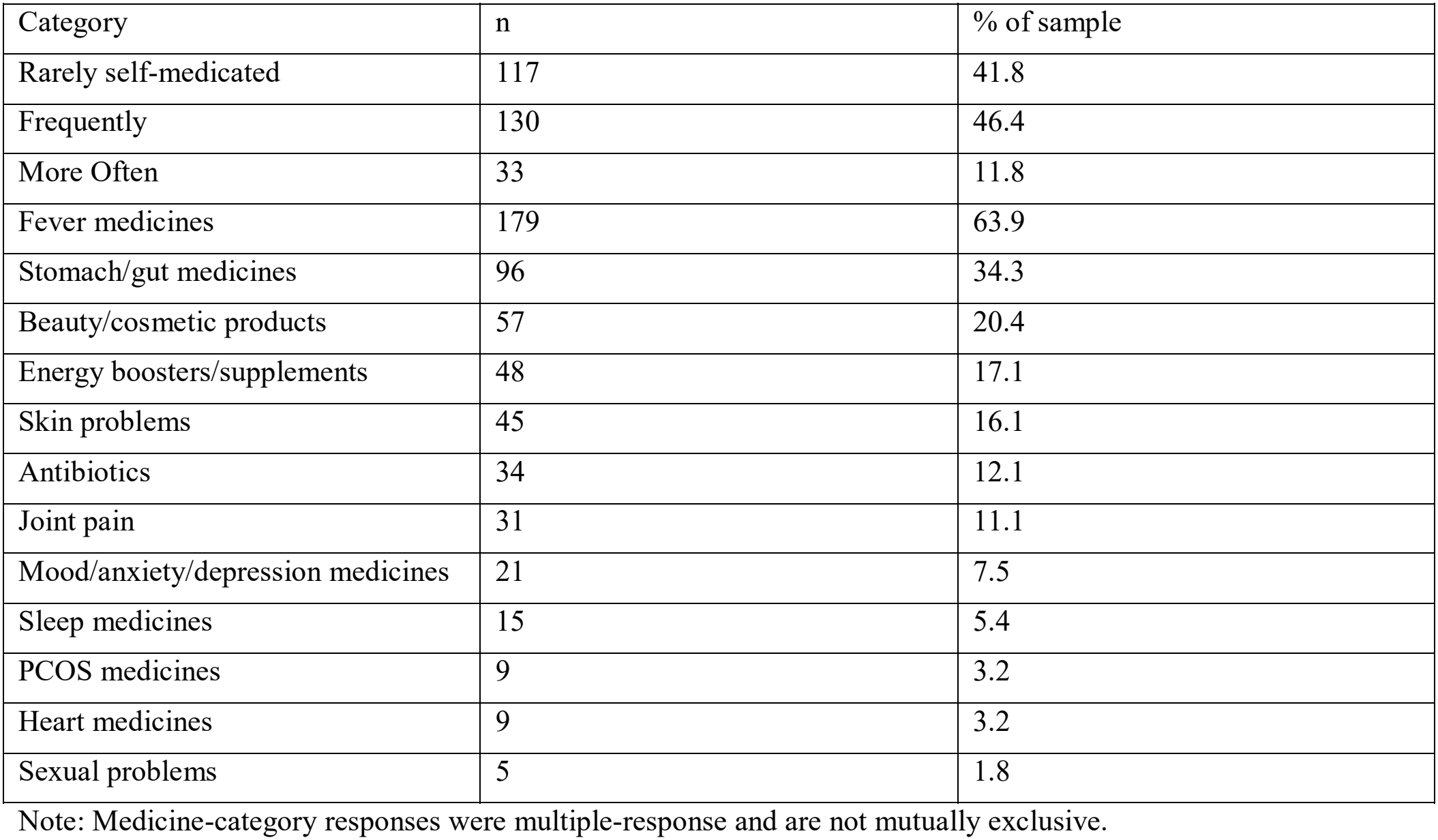
Self-medication frequency and reported medicine categories.

| Category | n | % of sample |
|---|---|---|
| Rarely self-medicated | 117 | 41.8 |
| Frequently | 130 | 46.4 |
| More Often | 33 | 11.8 |
| Fever medicines | 179 | 63.9 |
| Stomach/gut medicines | 96 | 34.3 |
| Beauty/cosmetic products | 57 | 20.4 |
| Energy boosters/supplements | 48 | 17.1 |
| Skin problems | 45 | 16.1 |
| Antibiotics | 34 | 12.1 |
| Joint pain | 31 | 11.1 |
| Mood/anxiety/depression medicines | 21 | 7.5 |
| Sleep medicines | 15 | 5.4 |
| PCOS medicines | 9 | 3.2 |
| Heart medicines | 9 | 3.2 |
| Sexual problems | 5 | 1.8 |

Note: Medicine-category responses were multiple-response and are not mutually exclusive.

**Table 3.**
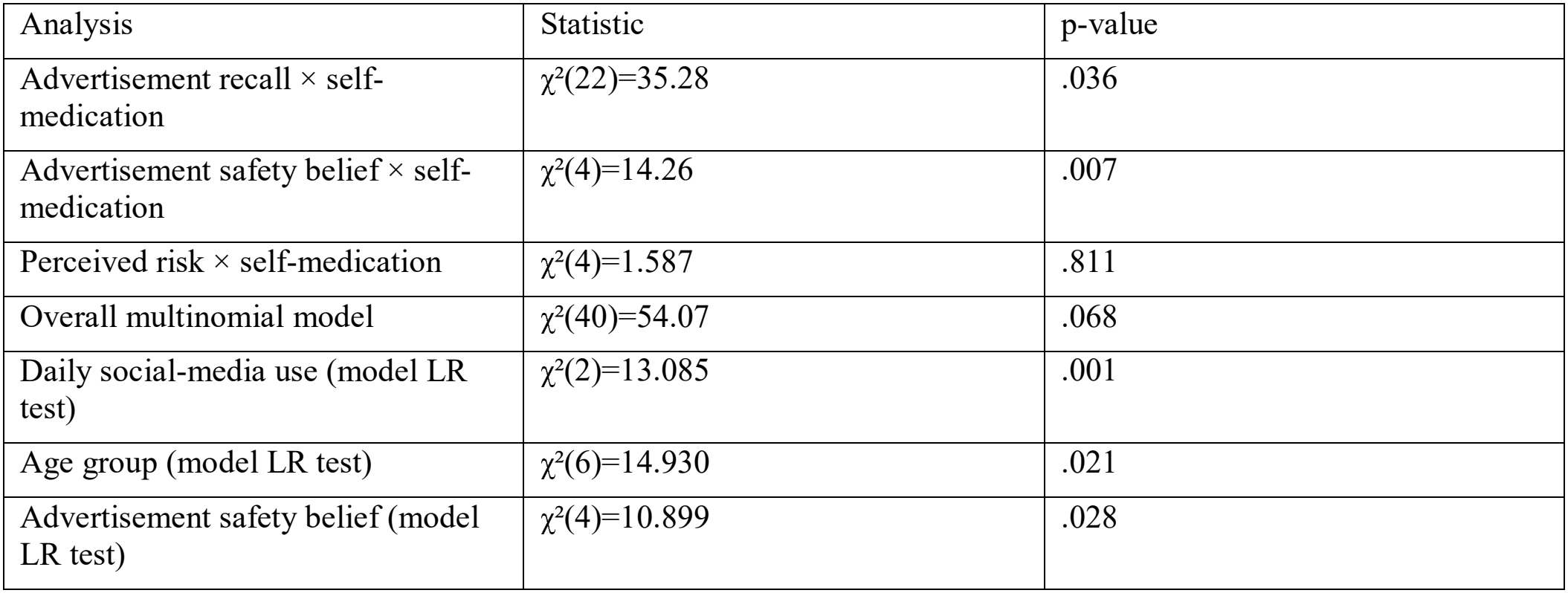
Key inferential findings.

| Analysis | Statistic | p-value |
|---|---|---|
| Advertisement recall × self-medication | χ²(22)=35.28 | .036 |
| Advertisement safety belief × self-medication | χ²(4)=14.26 | .007 |
| Perceived risk × self-medication | χ²(4)=1.587 | .811 |
| Overall multinomial model | χ²(40)=54.07 | .068 |
| Daily social-media use (model LR test) | χ²(2)=13.085 | .001 |
| Age group (model LR test) | χ²(6)=14.930 | .021 |
| Advertisement safety belief (model LR test) | χ²(4)=10.899 | .028 |

## 4. Discussion

This sample reported self-medication with some frequency, with fever and gastrointestinal medicines among the commonly reported categories. Instagram was the most frequently selected platform for social-media exposure. Beliefs about the safety of medicines presented in advertisements/posts were associated with self-medication frequency, suggesting that attitudes toward promotional content may be relevant to medicine-use behaviour.

Daily social-media-use duration and age group also showed likelihood-ratio associations in the multinomial model. However, the overall model test was not statistically significant at the prespecified .05 threshold; individual predictor findings should therefore be interpreted cautiously. The very wide confidence interval for the reported age comparison suggests sparse data in some categories and limits precision.

Advertisement recall differed across self-medication frequency groups, but the cross-sectional design does not establish that exposure preceded or caused self-medication. The absence of an association between perceived risk and self-medication frequency should not be interpreted as proof that risk knowledge is irrelevant. Practical digital-health-literacy initiatives could encourage users to evaluate sources, identify sponsored content, check medicine claims, and consult qualified healthcare professionals. Future studies should use larger, more representative samples and validated exposure measures, and consider longitudinal designs.

## 5. Limitations

The survey required smartphone and internet access, potentially excluding digitally disconnected individuals. The sample was predominantly young and university educated, limiting generalisability. Self-reported medicine use and a six-month recall period may introduce recall and social-desirability bias. The cross-sectional design cannot establish temporality or causality. Duplicate-response controls were limited; the thesis reports manual screening. Some contingency-table analyses had small expected cell counts. The overall multinomial regression model did not meet the .05 significance threshold; selected coefficient estimates, particularly the age comparison, had wide confidence intervals.

## 6. Conclusion

Self-medication was frequently reported in this sample of adults in Punjab, Pakistan. Medicine-advertisement safety beliefs, daily social-media-use duration, and age group showed associations with self-medication frequency in analyses; however, the overall multinomial model was not statistically significant, and the cross-sectional design does not support causal inference. Larger, more representative studies are needed.

## Supporting information

Supplementary Questionnaire

## Data Availability

The data generated and analyzed during this study are available from the corresponding author upon reasonable request.

## 7. Declarations

Ethics approval and consent to participate: Ethical approval was obtained from the Research Ethics and Support Committee (RESC), University of Management and Technology, Lahore, Pakistan (Reference No. RE-42-2026). Informed consent was obtained from all participants before data collection.

## Conflicts of interest

The authors declare no conflicts of interest.

### Data availability

The anonymised dataset generated and analysed during this study is available from the corresponding author upon reasonable request.

### Author contributions

Ayesha Ashraf: Conceptualization, methodology, investigation, data collection, data analysis, interpretation of results, and manuscript writing.

Faiza Khalid, Mian Salman, Muhammad Sharjeel, Muhammad Zohaib Qasim, and Muhammad Abu Bakar Saddique: Questionnaire development, data collection, data organization, interpretation of results, and manuscript review and editing.

All authors contributed to the research, reviewed the manuscript, and approved the final version for publication.

## Authors

Ayesha Ashraf, Faiza Khalid, Mian Salman, Muhammad Sharjeel, Muhammad Zohaib Qasim, Muhammad Abu Bakar Saddique

## Figures

**Figure 1.**
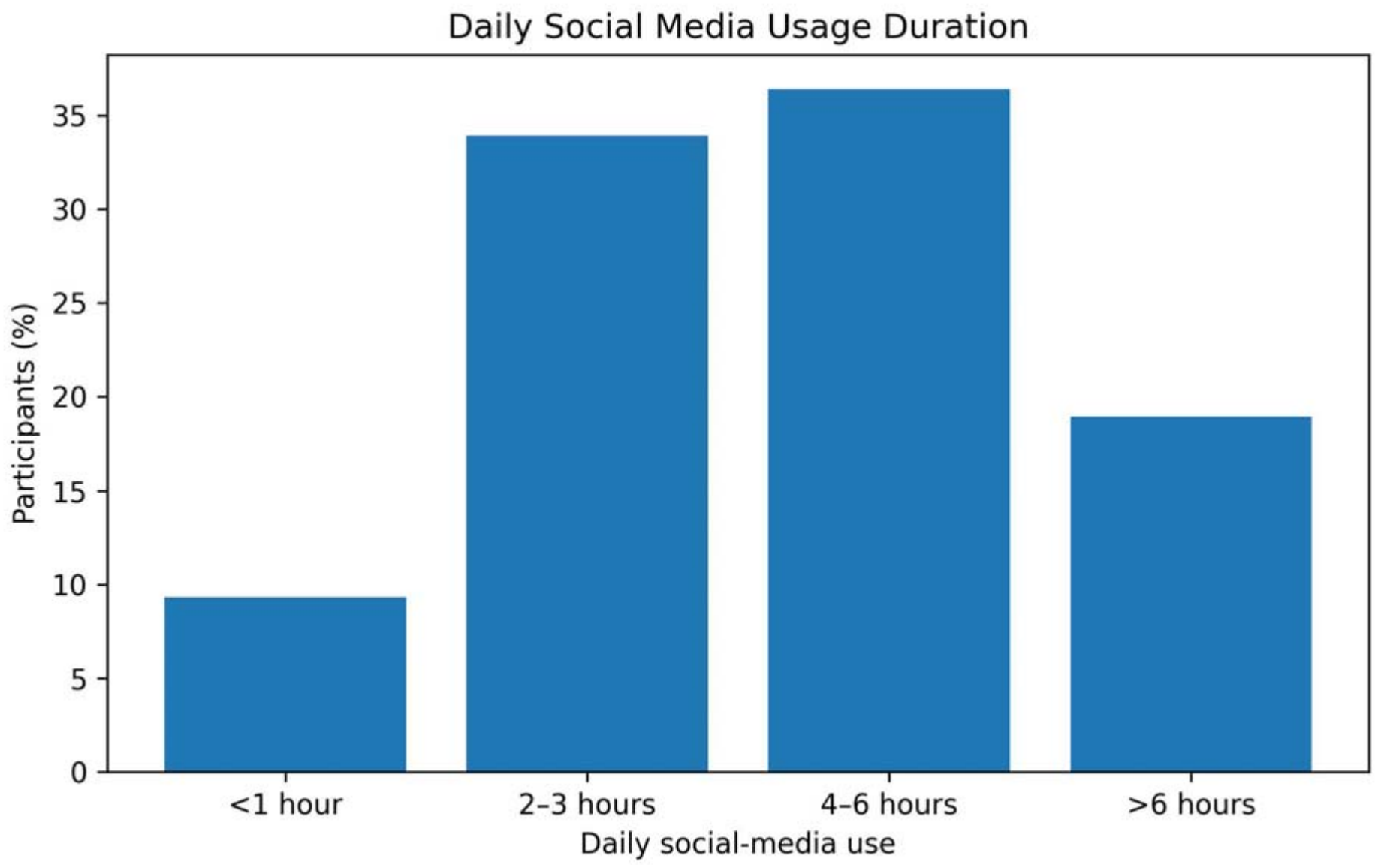
Daily social-media usage duration among participants.

**Figure 2.**
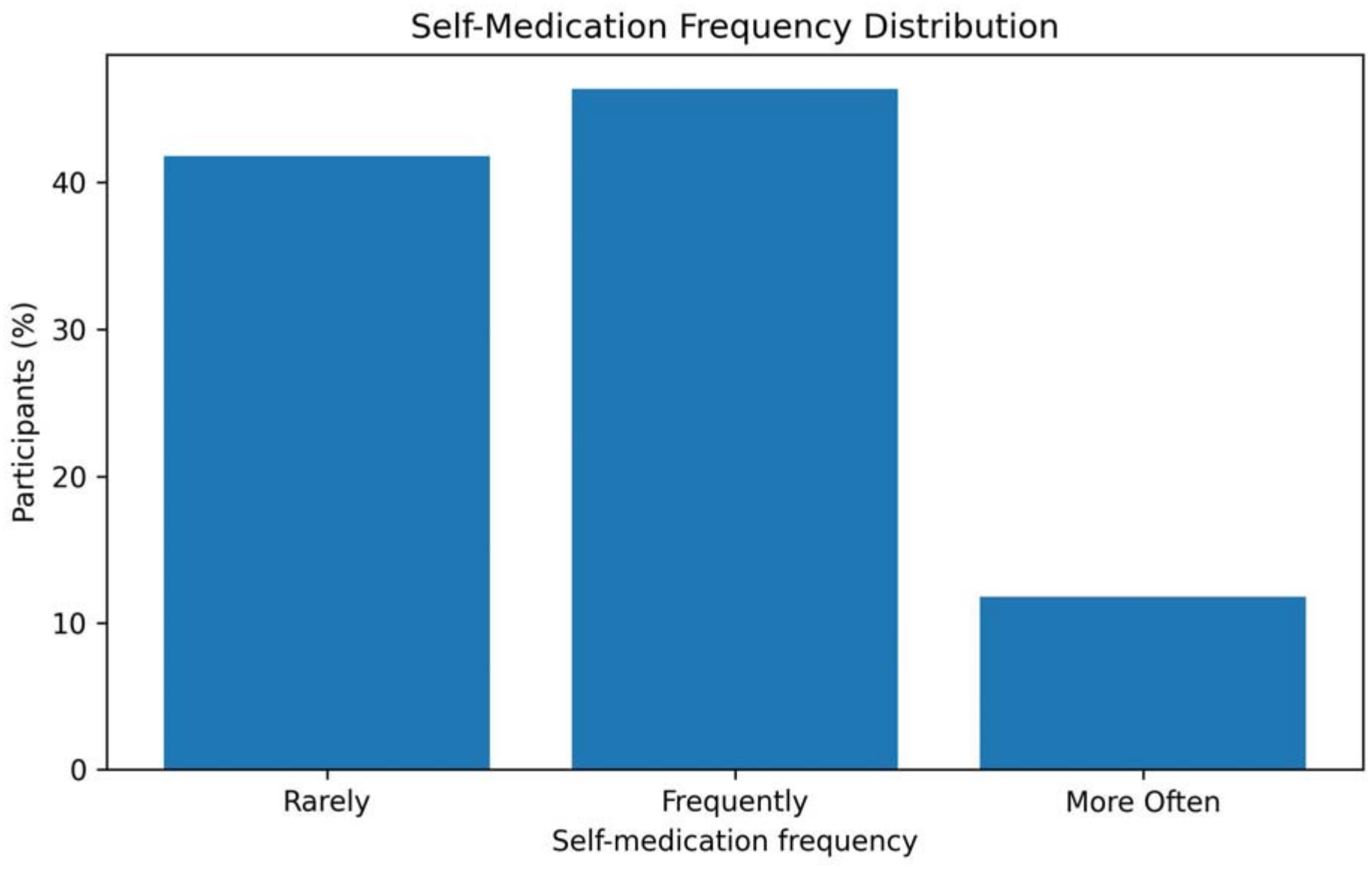
Self-medication frequency distribution among participants.

**Figure 3.**
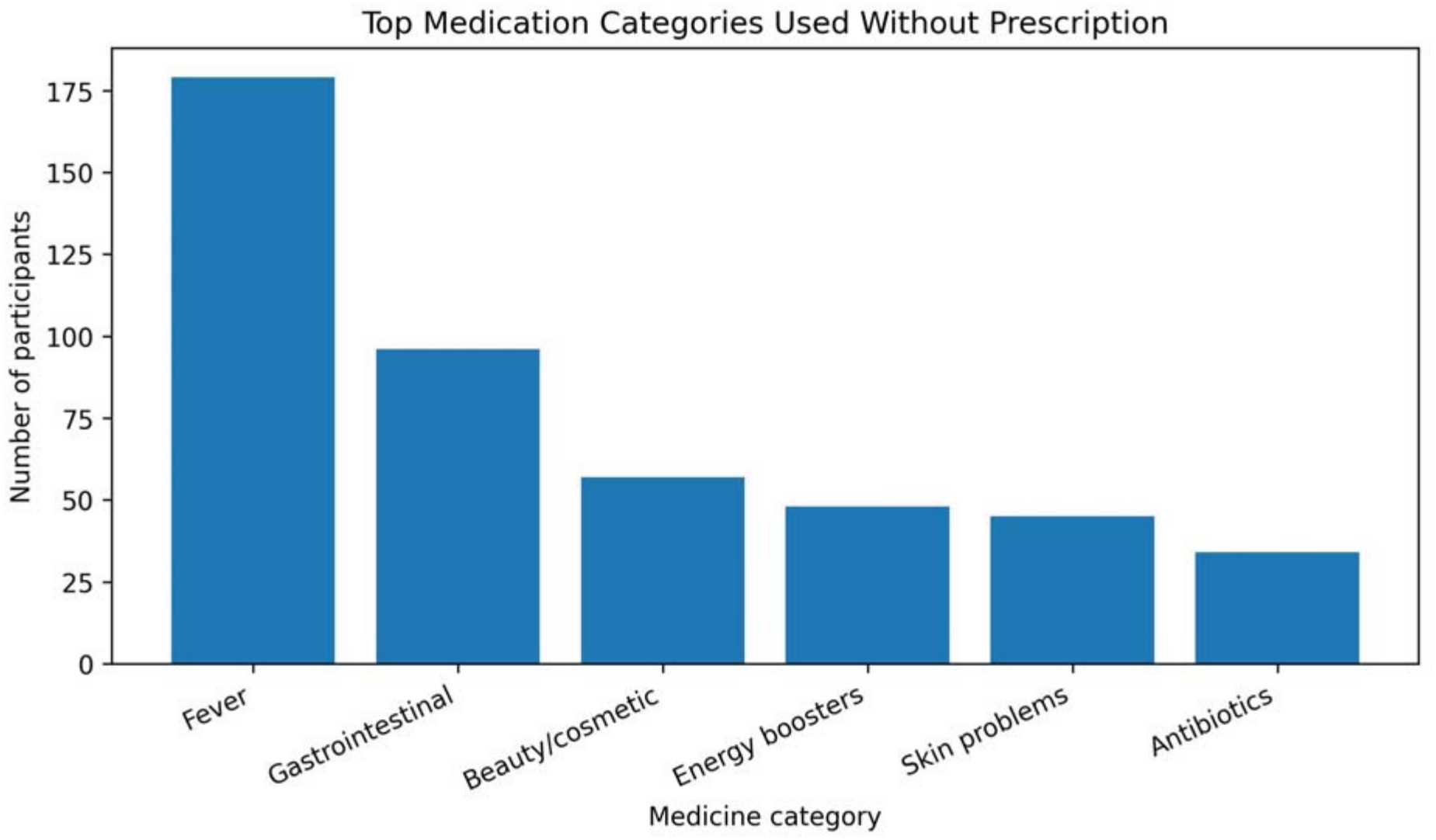
Top medication categories used without prescription. Multiple-response categories were not mutually exclusive.

## References

Alhabash, S., Dong, Y., Moureaud, C., Muraro, I. S., & Hertig, J. B. (2022). Effects of fear and humor appeals in public service announcements (PSAs) on intentions to purchase medications via social media. International Journal of Environmental Research and Public Health, 19(19), 12340.

Alhuthail, A., Aloraini, B., Alhuthail, I., Alrashidi, S. M., Alotibi, R. M., Alrashidi, S., & Alotibi, R. (2025). Attitudes and awareness regarding health information sought on social media by the Saudi population: A cross-sectional study. Cureus, 17(1).

Aziz, M. M., Masood, I., Yousaf, M., Saleem, H., Ye, D., & Fang, Y. (2018). Pattern of medication selling and self-medication practices: A study from Punjab, Pakistan. PLoS ONE, 13(3), e0194240.

Baig, I., Zia, A. Z. A., & Awais, M. (2025). Social Media Use for Health-Related Information and Self-Medication Practices among Youth: Cognitive Dissonance Perspective. Lahore Institute for Research and Analysis Journal, 3, 13–26.

Cotobal Calvo, E. M., Bocchino, A., Mata-Pérez, C., Cruz-Barrientos, A., Naranjo-Márquez, M., & Palazón-Fernández, J. L. (2024). Content validation of the self-medication scale and trust in online resources: Deepening digital access to health. Nursing Reports, 14(3), 1897–1905.

Do Nascimento, I. J. B., Pizarro, A. B., Almeida, J. M., Azzopardi-Muscat, N., Gonçalves, M. A., Björklund, M., & Novillo-Ortiz, D. (2022). Infodemics and health misinformation: a systematic review of reviews. Bulletin of the World Health Organization, 100(9), 544.

Ghimire, P., Pant, P., Khatiwada, S., Ranjit, S., Malla, S., & Pandey, S. (2023). Self-medication practice in Kathmandu Metropolitan City: A cross-sectional study. SAGE Open Medicine, 11, 20503121231158966.

Hamza, A., Yonghong, D., & Ullah, I. (2025). Dynamic shifts in social media usage in Pakistan: A comparative analysis across pre-, during-, and post-COVID-19 periods. Journalism and Media, 6(2), 59.

Ittefaq, M., Seo, H., Abwao, M., & Baines, A. (2022). Social media use for health, cultural characteristics, and demographics: A survey of Pakistani millennials. Digital Health, 8, 20552076221089454.

Kemp, S. (2025). Digital 2025: Pakistan. DataReportal. https://datareportal.com/reports/digital-2025-pakistan

Mahdi, S., Akram, W., Mumtaz, Z., Tariq, A., & Akhtar, A. (2026). AI Chatbots and Online Health Information Use in Self-Medication Among University Students in Lahore, Pakistan: A Cross-Sectional Study. Journal of Health, Wellness and Community Research, 1–10.

Mor, J., Kaur, T., Menkes, D. B., Peter, E., & Grundy, Q. (2024). Pharmaceutical industry promotional activities on social media: a scoping review. Journal of Pharmaceutical Health Services Research, 15(4), rmae022.

Nazari, J., Ghafoury, R., Chezani-Sharahi, N., Moradzadeh, R., & Naghshbandi, M. (2024). Prevalence and determinants of self-medication consumption of antibiotics in adults in Iran: a population based cross-sectional study, 2019–2020. Frontiers in Public Health, 12. 10.3389/fpubh.2024.1502074

Ocan, M., Obuku, E. A., Bwanga, F., Akena, D., Richard, S., Ogwal-Okeng, J., & Obua, C. (2015). Household antimicrobial self-medication: a systematic review and meta-analysis of the burden, risk factors and outcomes in developing countries. BMC Public Health, 15(1), 742.

Ogasawara, H., Japan, D. C. S., & Indonesia, M. (2000). WHO guidelines for the regulatory assessment of medicinal products for use in self-medication. WHO Drug Information, 14(1), 6.

Penagos-Corzo, J. C., Ortiz-Barrero, M. J., Hernández-Ramírez, R., Ochoa-Ramírez, Y., González Ehlinger, R., & Pérez-Acosta, A. M. (2024). Development and psychometric properties of a self-medication behavior inventory. Frontiers in Psychology, 15, 1366284.

Rathod, P., Sharma, S., Ukey, U., Sonpimpale, B., Ughade, S., Narlawar, U., Gaikwad, S., Nair, P., Masram, P., & Pandey, S. (2023). Prevalence, pattern, and reasons for self-medication: a community-based cross-sectional study from central India. Cureus, 15(1).

Saifullah, M., Sharafi, S., & Ghanei Gheshlagh, R. (2025). Prevalence and patterns of self-medication in Pakistan: a systematic review and meta-analysis. BMC Public Health.

Sarhan, N., Schaalan, M. F., & El-Sheikh, A. A. K. (2025). Patterns and predictors of self-medication behavior of weight loss medications: a cross-sectional analysis of social media influence and role of pharmacist intervention. Frontiers in Pharmacology, 16, 1606566. 10.3389/fphar.2025.1606566

Taba, M., Allen, T. B., Caldwell, P. H., Skinner, S. R., Kang, M., McCaffery, K., & Scott, K. M. (2022). Adolescents’ self-efficacy and digital health literacy: a cross-sectional mixed methods study. BMC Public Health, 22(1), 1223.

Ventola, C. L. (2014). Social media and health care professionals: benefits, risks, and best practices. Pharmacy and Therapeutics, 39(7), 491.

Willis, E., Friedel, K., Heisten, M., Pickett, M., & Bhowmick, A. (2023). Communicating health literacy on prescription medications on social media: in-depth interviews with “patient influencers”. Journal of Medical Internet Research, 25, e41867.

