## Supplementary Tables for "Association between Social Media Exposure and Self-Medication Practices: Public Health Implications in Punjab, Pakistan"

### SOCIO-DEMOGRAPHIC CHARACTERISTICS

A total of 280 completed responses were analyzed. The sociodemographic profile of the sample is detailed in Table 1 below:

| Variable | Category | n | % |
|---|---|---|---|
| **Age** | 18–25 yrs | 206 | 73.6% |
| | 25–35 yrs | 45 | 16.1% |
| | 35–45 yrs | 22 | 7.9% |
| | 45–60 yrs | 7 | 2.5% |
| **Gender** | Male | 138 | 49.3% |
| | Female | 142 | 50.7% |
| **Education** | University+ | 199 | 71.1% |
| | College | 62 | 22.1% |
| **Occupation** | Student/Dependent | 158 | 56.4% |
| | Private sector | 47 | 16.8% |

*Table 1. Socio-demographic characteristics of study participants (N=280)*

### KEY FINDINGS SUMMARY

**58.2%**

Self-medicated frequently or more often

**63.9%**

Used antipyretics without prescription

**36.4%**

Used social media 4–6 hrs/day

**50.0%**

Followed medical journey accounts

**DATA VISUALIZATIONS & DISTRIBUTIONS**

To provide complete reference and editing capabilities, the figures below include both the graphical representations and their underlying empirical data distributions.

***Figure 1. Self-medication frequency distribution (N=280)***

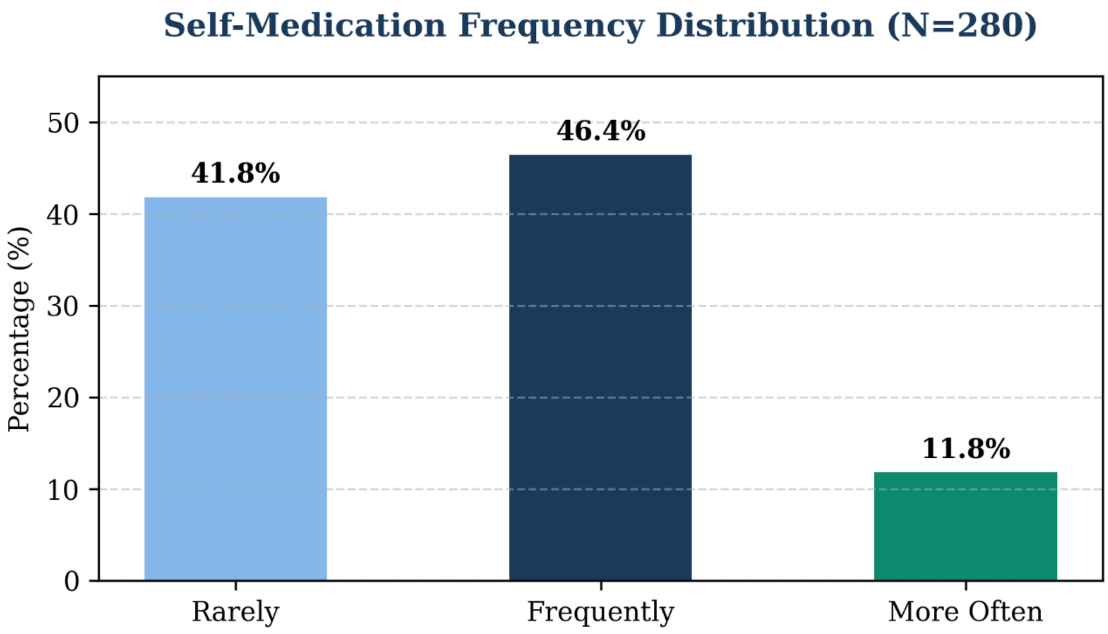


| Self-Medication Frequency | Percentage (%) | Estimated Cases (n) |
|---|---|---|
| Rarely | 41.8% | 117 |
| Frequently | 46.4% | 130 |
| More Often | 11.8% | 33 |
| Total | 100.0% | 280 |

***Figure 2. Top medication categories used without prescription (multiple response, cases)***

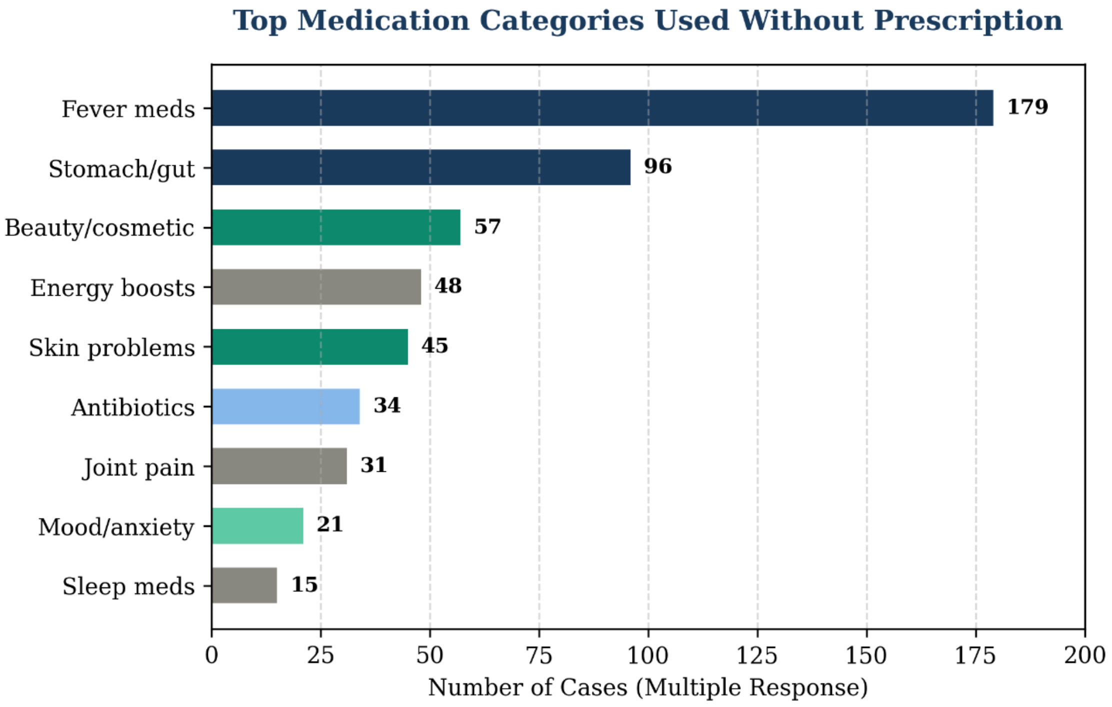


| Medication Category | Number of Cases | Percentage of Sample (N=280) |
|---|---|---|
| Fever medications (Antipyretics) | 179 | 63.9% |
| Stomach / Gut medications | 96 | 34.3% |
| Beauty / Cosmetic products | 57 | 20.4% |
| Energy boosters / Vitamins | 48 | 17.1% |
| Skin problem treatments | 45 | 16.1% |
| Antibiotics | 34 | 12.1% |
| Joint pain analgesics | 31 | 11.1% |
| Mood / Anxiety medications | 21 | 7.5% |
| Sleep medications | 15 | 5.4% |

***Figure 3. Daily social media usage duration (N=280)***

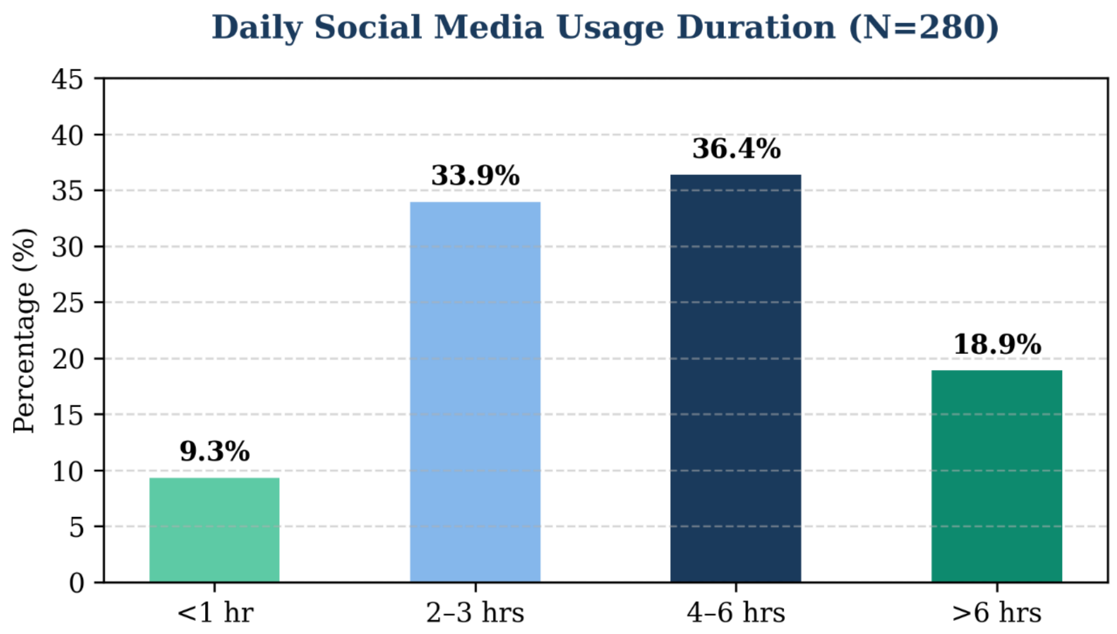


| Daily Social Media Usage | Percentage (%) | Number of Cases (n) |
|---|---|---|
| Less than 1 hour | 9.3% | 26 |
| 2–3 hours | 33.9% | 95 |
| 4–6 hours | 36.4% | 102 |
| More than 6 hours | 18.9% | 57 |
| Total | 100.0% | 280 |

***Figure 4. Platforms for pharmaceutical advertisement recall (multiple response)***

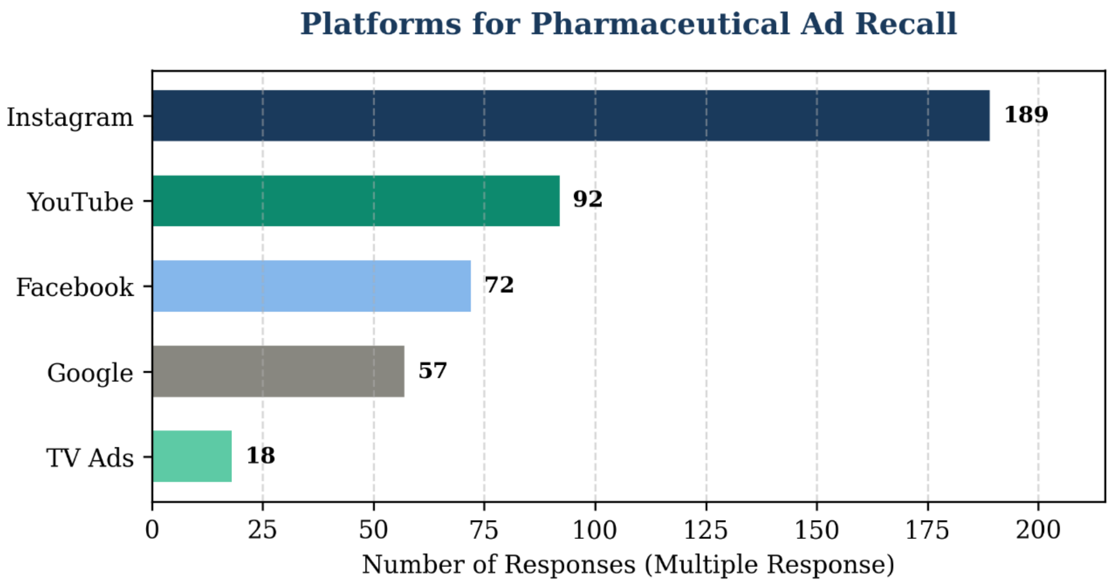


| Platform | Number of Responses | Rank |
|---|---|---|
| Instagram | 189 | 1 |
| YouTube | 92 | 2 |
| Facebook | 72 | 3 |
| Google Search | 57 | 4 |
| Television Ads | 18 | 5 |

**STATISTICAL ANALYSIS RESULTS**

Inferential statistical analyses were performed to evaluate the independent predictors of self-medication frequency. Table 2 presents the Multinomial Logistic Regression (NOMREG) likelihood ratio tests:

| Predictor Variable | Likelihood Ratio $\chi^2$ | df | p-value | Statistical Significance |
|---|---|---|---|---|
| Daily SM usage (SMtime) | 13.085 | 2 | **.001**** | **Significant** |
| Age group | 14.930 | 6 | **.021*** | **Significant** |

| | | | | |
|---|---|---|---|---|
| Medicines in ads safe (SafeAds) | 10.899 | 4 | **.028*** | **Significant** |
| SM made self-treatment easier | 3.071 | 2 | .215 | Non-sig. |
| SM influence on medication | 2.247 | 2 | .325 | Non-sig. |
| Risk awareness (MedRisk) | 0.026 | 2 | .987 | Non-sig. |
| Gender | 2.008 | 2 | .366 | Non-sig. |
| Qualification | 1.654 | 4 | .799 | Non-sig. |

*Table 2. NOMREG Likelihood Ratio Tests — predictors of self-medication frequency (reference category: More Often). Nagelkerke $R^2 = .205$. *p<.05 **p<.01*

Furthermore, Chi-square tests of independence were executed between attitudinal variables and self-medication frequency, as shown in Table 3:

| Attitudinal Variable | $\chi^2$ Value | df | p-value | Statistically Significant? |
|---|---|---|---|---|
| Medicines in ads are safe (SafeAds) | 14.260 | 4 | **.007*** | **Yes (Fisher's p=.006)** |
| SM can replace doctor | 5.083 | 4 | .279 | No |
| Lack info about drugs on SM | 5.682 | 4 | .224 | No |
| Medication use from SM has risks | 1.587 | 4 | .811 | No |
| Trust medical advice from SM | 4.170 | 4 | .383 | No |

*Table 3. Chi-square tests of independence: attitudinal variables vs self-medication frequency.*
