## Supplementary Questionnaire for "Association between Social Media Exposure and Self-Medication Practices: Public Health Implications in Punjab, Pakistan"

This questionnaire is part of a research study aimed at understanding the impact of social media on self-medication practices. Your responses will help assess how online information influences medicine use and health decisions. The questionnaire will take about 2-5 minutes to complete.

* Indicates required question

1. Please select your language براہ کرم اپنی زبان منتخب کریں *

*Mark only one oval.*

- English *Skip to question 18*
- اردو *Skip to question 2*

#### سوشل میڈیا کی نمائش اور خود ادویات کے طریقوں کے درمیان تعلق: پنجاب، پاکستان میں صحت عامہ اثرا

سوشل میڈیا کی نمائش اور خود ادویات کے طریقوں کے درمیان تعلق: پنجاب، پاکستان میں صحت عامہ کے اثرات
ہم ایک تحقیقی مطالعہ کر رہے ہیں جس کا مقصد یہ جاننا ہے کہ لوگ ادویات کے بارے میں معلومات حاصل کرنے کے لیے سوشل میڈیا کو کس حد تک استعمال کرتے ہیں اور یہ ان کے خود علاج کے رویے کو کیسے متاثر کرتا ہے۔
آپ کی اس تحقیق میں شرکت مکمل طور پر رضاکارانہ ہے اور آپ کی فراہم کردہ تمام معلومات کو خفیہ رکھا جائے گا اور صرف تحقیقی مقاصد کے لیے استعمال کیا جائے گا۔
اس سوالنامے کو مکمل کرنے میں تقریباً 2 سے 5 منٹ لگیں گے

2. آپ رضاکارانہ طور پر اس تحقیق میں حصہ لینے پر رضامند ہیں۔ *

*Check all that apply.*

- [ ] میں رضامند ہوں

#### ذاتی معلومات

3. 1. آپ کی عمر کیا ہے؟ *

*Mark only one oval.*

- 18–25 سال
- 25–35 سال
- 35–45 سال
- 45–60 سال

4. 2. آپ کی جنس کیا ہے؟ *

*Check all that apply.*

- [ ] مرد
- [ ] عورت
- [ ] دیگر

5. 3. آپ کی تعلیمی قابلیت کیا ہے؟ *

*Mark only one oval.*

- اسکول
- کالج
- یونیورسٹی یا اس سے زیادہ

6. 4. آپ کا پیشہ کیا ہے؟ *

*Mark only one oval.*

- نجی شعبہ
- خود روزگار
- بے روزگار
- طالب علم / زیر کفالت
- سرکاری ملازم

#### خود علاج

7. 5. کیا آپ نے کبھی ڈاکٹر کے نسخے کے بغیر دوا استعمال کی ہے؟ *

*Mark only one oval.*

- کبھی کبھار
- اکثر
- بہت زیادہ

8. 6 . آپ عام طور پر خود علاج کیوں کرتے ہیں؟ (تمام منتخب کریں جو موزوں ہوں) *

*Check all that apply.*

- [ ] معمولی بیماری
- [ ] مصروف شیڈول یا روزمرہ مصروفیات
- [ ] خود علاج کم خرچ ہوتا ہے
- [ ] ہسپتال یا کلینک میں زیادہ انتظار
- [ ] پہلے سے دوا کا تجربہ
- [ ] Other: ____________

9. 7. آپ روزانہ سوشل میڈیا پر تقریباً کتنا وقت گزارتے ہیں؟ *

*Mark only one oval.*

- 1 گھنٹے سے کم
- 2–3 گھنٹے
- 4–6 گھنٹے
- 6 گھنٹے سے زیادہ

10. آپ کون سا سوشل میڈیا پلیٹ فارم زیادہ استعمال کرتے ہیں؟ (تمام منتخب کریں جو موزوں ہوں) . 8 *

*Check all that apply.*

- [ ] انسٹاگرام
- [ ] ٹی وی اشتہارات
- [ ] گوگل
- [ ] فیس بک
- [ ] یوٹیوب
- [ ] Other: ______

11. کیا آپ سوشل میڈیا پر ایسے لوگوں کو فالو کرتے ہیں جو اپنی میڈیکل جرنی یا علاج کا تجربہ شیئر کرتے ہیں؟9. *

*Mark only one oval.*

- ہاں
- نہیں

#### میڈیا کے اثر سے خود علاج

12. سوشل میڈیا کے اثر سے آپ عام طور پر کون سی ادویات استعمال کرتے ہیں؟ ( تمام منتخب کریں جو موزوں ہوں) 10. *

*Check all that apply.*

- [ ] بخار کی ادویات
- [ ] معدہ یا ہاضمے کی ادویات
- [ ] دل کی بیماریوں کی ادویات
- [ ] مختلف انفیکشن کے لیے اینٹی بائیوٹکس
- [ ] وزن کم یا زیادہ کرنے کی ادویات
- [ ] ذہنی دباؤ یا ڈپریشن کی ادویات
- [ ] خوبصورتی یا کاسمیٹک مصنوعات
- [ ] جوڑوں کے درد کی ادویات
- [ ] توانائی بڑھانے والے سپلیمنٹس
- [ ] جلد کی بیماریوں کی ادویات
- [ ] نیند کی ادویات
- [ ] جنسی مسائل کی ادویات
- [ ] ماہواری کے مسائل کی ادویات
- [ ] PCOS(polycystic ovarian syndrome)
- [ ] Other: ____________________

13. 11. سوشل میڈیا پر آپ نے زیادہ تر کس قسم کے اشتہارات دیکھے ہیں؟ ( تمام منتخب کریں جو موزوں ہوں ) *

*Check all that apply.*

- [ ] بالوں کے مسائل
- [ ] وزن بڑھانا
- [ ] وزن کم کرنا
- [ ] ماہواری کے مسائل
- [ ] ذیابیطس
- [ ] قد بڑھانا
- [ ] جوڑوں کا درد
- [ ] بلڈ پریشر
- [ ] جنسی مسائل
- [ ] Other: ____________

#### سوشل میڈیا کی بنیاد پر ادویات کے استعمال کے بارے میں معلومات کا جائزہ

14. *

*Mark only one oval per row.*

| | متفق | غیر متفق | غیر جانبدار |
|---|---|---|---|
| کیا آپ سمجھتے ہیں کہ سوشل میڈیا پر ادویات کے بارے میں معلومات کی کمی ہوتی ہے؟ 12. | ⬭ | ⬭ | ⬭ |
| کیا آپ سمجھتے ہیں کہ سوشل میڈیا کی بنیاد پر دوا استعمال کرنے میں خطرات ہو سکتے ہیں؟ 13. | ⬭ | ⬭ | ⬭ |

15. 14: کیا آپ کے خیال میں سوشل میڈیا ڈاکٹر سے مشورہ کرنے کا متبادل ہو سکتا ہے؟ *

*Mark only one oval.*

- متفق
- غیر متفق
- غیر جانبدار

16. 15: سوشل میڈیا آپ کے ادویات کے استعمال پر کتنا اثر انداز ہوتا ہے؟

*Mark only one oval.*

- کبھی کبھار
- اکثر
- بہت زیادہ

***درج ذیل بیانات کے بارے میں اپنی رائے دیں***

17. *

*Mark only one oval per row.*

| | متفق | غیر جانبدار | متفق نہیں | بالکل متفق نہیں |
|---|---|---|---|---|
| میں سوشل میڈیا پر لوگوں کے طبی مشوروں پر اعتماد کرتا ہوں۔16. | ⬭ | ⬭ | ⬭ | ⬭ |
| سوشل میڈیا مجھے ڈاکٹر کے بغیر اپنا علاج کرنا آسان بناتا ہے۔17. | ⬭ | ⬭ | ⬭ | ⬭ |
| مجھے لگتا ہے کہ اشتہارات یا پوسٹس میں دکھائی جانے والی ادویات استعمال کے لیے محفوظ ہیں۔18. | ⬭ | ⬭ | ⬭ | ⬭ |

### Association Between Social Media Exposure and Self-Medication Practices: Public Health Implications in Punjab, Pakistan

This questionnaire is part of a research study aimed at understanding the impact of social media on self-medication practices. Your responses will help assess how online information influences medicine use and health decisions. All information provided will remain confidential and will be used for academic purposes only.

18. **You voluntarily agree to participate in this research.** *

*Check all that apply.*

- [ ] I Agree

#### Demographics and Health Background

19. Q1: What is your age? *

*Mark only one oval.*

- ( ) 18-25 years.
- ( ) 25-35 years.
- ( ) 35-45 years.
- ( ) 45-60 years.

20. Q2: What is your gender? *

*Mark only one oval.*

- ( ) Male.
- ( ) Female.
- ( ) Other

21. Q3 What is your highest qualification?

*Mark only one oval.*

- School.
- College .
- University and above.

22. Q4: What is your Occupation? *

*Mark only one oval.*

- Private sector.
- Self employed.
- Unemployed .
- Student / dependent.
- Government employee

**Self Medication History:**

23. Q5: Have you used medication without a doctor prescription? *

*Mark only one oval.*

- Rarely.
- Frequently.
- More often.

24. Q6: What is your reason for self-medication mostly? (Select all that apply). *

*Check all that apply.*

- [ ] Minor illness.
- [ ] Busy schedule / routine.
- [ ] Self-medication is inexpensive.
- [ ] Long wait at the health facility.
- [ ] Prior experience with medication.
- [ ] Other: ____________________

25. Q7: What is your approximate daily time on social media usage? *

*Mark only one oval.*

- ( ) Less than 1 hour
- ( ) 2-3 hours
- ( ) 4-6 hours
- ( ) More than 6 hours

26. Q8: Which social media platform do you use frequently? (Select all that apply) *

*Check all that apply.*

- [ ] Instagram
- [ ] TV Advertisement
- [ ] Google
- [ ] Facebook
- [ ] YouTube
- [ ] Other: ____________________

27. Q9: Do you follow people on social media who share their own "medical journey"? *

*Mark only one oval.*

- Yes
- No

***Self-medication under media influence:***

28. Q10: Which class of drugs do you use commonly under media influence? (Select all that apply). *

*Check all that apply.*

- [ ] Medications for fever.
- [ ] Medications for stomach/gut issues.
- [ ] Medications for heart problems.
- [ ] Antibiotics for various infections.
- [ ] Medications for weight management.
- [ ] Medications for mood changes (anxiety or depression).
- [ ] Beauty and cosmetic products.
- [ ] Medications for joint pain.
- [ ] Energy booster/ supplement.
- [ ] Medications for skin problems.
- [ ] Medications for sleep.
- [ ] Medications for sexual problems.
- [ ] Medications for menstrual problems.
- [ ] PCOS (polycystic ovarian syndrome)
- [ ] Other: ____________________

29. Q11: List of advertisements you have mostly seen on social media? (select all that apply) *

*Check all that apply.*

- [ ] Hair problem.
- [ ] Weight gain.
- [ ] Weight loss .
- [ ] Menstrual problems.
- [ ] Diabetes.
- [ ] Height gain.
- [ ] Joint pain.
- [ ] Blood pressure.
- [ ] Sexual problem.
- [ ] Skin issues.
- [ ] Other: ____________________

***Evaluation of knowledge on drug use based on social media:***

30. Q12: Do you think there is a lack of information about drugs on social media? *

*Mark only one oval.*

- ( ) Agree
- ( ) Disagree
- ( ) Neutral

31. Q13: . Do you think use of medication for health problems on social media have risks? *

*Mark only one oval.*

- Agree
- Disagree
- Neutral

32. Q14: Do you think social media can be a substitute for consulting a doctor? *

*Mark only one oval.*

- Agree
- Disagree
- Neutral

33. Q15: How much social media influences your medication usage? *

*Mark only one oval.*

- Rarely.
- More often.
- frequently.

***Rate your level of agreement with the following:***

34. Statement *

*Mark only one oval per row.*

| | Strongly disagree | Disagree | Agree |
|---|---|---|---|
| **Q16: I trust medical advice from people on social media** | ⬭ | ⬭ | ⬭ |
| **Q17: Social media makes it easier to treat myself without a doctor.** | ⬭ | ⬭ | ⬭ |
| **Q18: I believe medication shown in the ads or posts is safe to use** | ⬭ | ⬭ | ⬭ |

***Thank you for your participation! آپ کی شرکت کا شکریہ!***

This content is neither created nor endorsed by Google.

Google Forms
